# Speaking in Tones: The role of lexical tones in Chinese-speaking Primary Progressive Aphasia

**DOI:** 10.1101/2025.10.24.25338751

**Authors:** Boon Lead Tee, Ta-Fu Chen, Lorinda Kwan-Chen Li-Ying, Raymond Y. Lo, Joshua Tsoh, Andrew Lung-Tat Chan, Adrian Wong, Chien Jung Lu, Yu Sun, Pei-Ning Wang, YiChen Lee, Isabel Elaine Allen, Yu-Chen Chuang, Yu-Wen Cheng, FangDa Leng, Yu Chen, Maria-Luisa Mandelli, Jessica de Leon, Jet Vonk, Maria Luisa Gorno-Tempini

**Author notes:** Correspondence to: Boon Lead Tee MD MSc Full address: University of California, San Francisco, Memory and Aging Center 675 Nelson Rising Lane, Suite 190, San Francisco 94158, California, United States.

## Abstract

Two-thirds of the world’s languages, including Mandarin and Cantonese, employ pitch variation to convey meaning (lexical tone). Existing diagnostic frameworks for primary progressive aphasia (PPA) have been developed for English speakers, and have not considered the impact of salient language-specific variations, such as tone. This study investigates lexical tone processing in Mandarin- and Cantonese-speaking individuals with PPA and examines their neural signatures using structural neuroimaging.

**Methods:** Seventy-eight native Chinese speakers (54 with PPA; 24 healthy controls) were assessed using the CLAP (Chinese Language Assessment for PPA) battery, a series of neuropsychological and linguistic tasks developed to characterize the linguistic features of Mandarin and Cantonese speakers with PPA. Lexical tone production was examined through repetition and reading of “tone-twister” phrases, as well as repetition of multicharacter phrases varying in articulatory features (place, manner, and tone). Tone perception and comprehension was assessed via identification, discrimination, and tone-word/picture matching tasks. Group differences were analyzed using nonparametric tests and generalized estimating equations, with ROC analyses determining diagnostic accuracy. Structural MRI data were acquired for 55 participants, and voxel-based morphometry (VBM) was used to examine the neural correlates of tone performance.

**Results:** Participants with nonfluent/agrammatic variant PPA (nfvPPA) showed marked impairments in lexical tone production (all p<0.001), with a disproportionately high rate of tonal relative to syllabic errors (p<0.001). In contrast, semantic variant PPA (svPPA) exhibited prominent deficits in three tone perception tasks (all p<0.001). Patients with logopenic variant PPA (lvPPA) showed relatively preserved tone production but a predominance of syllabic errors (p<0.001), suggesting underlying phonological deficits. Lexical tone production tasks demonstrated strong discrimination of nfvPPA (AUC= 0.702-0.907). In contrast, three tone perception tasks exhibited high sensitivity for detecting svPPA (90.9-100%), though specificity was modest (37-63%). Neural correlate analyses revealed that tone production deficits were associated with reduced grey matter volume in the left inferior frontal gyrus, insula, and temporal cortex, whereas tone perception performance correlated with atrophy in the left superior and middle temporal gyri, temporal pole, and orbitofrontal regions.

**Discussion:** Lexical tone processing is differentially impaired across PPA subtypes, with tone production and perception deficits mapping onto distinct neural substrates. These findings underscore the necessity of developing language-specific diagnostic approaches for tonal language speakers and call into question the cross-cultural applicability of current PPA diagnostic strategies, which have been largely shaped by Indo-European language frameworks.

## Introduction

Nearly all human languages, in their spoken forms, construct words based on fundamental units of sound (phonemes) which typically consist of consonants and/or vowels sounds. These phonemic building blocks are frequently organized into syllables, which are then merged to form words.^1^ However, around 60-70% of the world’s living spoken languages also utilize pitch variation at the word level, also known as lexical tone, to encode semantic, morphological, and grammatical meanings.^2,3^ These languages are commonly described as tonal languages and they are largely prevalent and concentrated in Asia, such as Mandarin, Cantonese, Thai and Vietnamese; in Africa, including Yoruba and Zulu; and in Central America, with languages like K’iche’ and other Mayan languages.^3^ (tonal language population worldwide) Despite the global prevalence of tone languages, neurolinguistic research has traditionally centered on segmental phonology (i.e., consonants and vowels), while the clinical significance of lexical tones to diagnose and monitor aphasia have received limited attention.^4^ Tonal languages such as Mandarin, Cantonese and Thai, rely heavily on tones to convey semantic meaning, therefore words composed of identical phonemes can differ entirely in meaning depending solely on their tonal contour. For example, the syllable */shi/*, when articulated with a high-level pitch (Tone 1; Supplementary Figure 1), can represent a word such as *shī* (詩, “poem”), whereas the same syllable produced with a high-falling pitch (Tone 4) changes meaning entirely, as in *shì* (是, “to be”). Another paradigmatic illustration of the intricacy and linguistic significance of tonal processing in Chinese is the one-syllable Mandarin poem “Lion-Eating Poet in the Stone Den” by Chinese linguist Yuen Ren Chao. The poem’s initial verse, “石室詩士施氏, 嗜獅, 誓食十獅,” is pronounced as /shi2 shi4 shi1 shi4 shi1 shi4, shi4 shi1, shi4 shi2 shi2 shi1/, which literally translates to “a poet named Shi lives in a stone room, ate a lion, and vowed to eat ten lions.” Although the verse consists entirely of the syllable/shi/, tonal variation serve as the primary means of differentiates the possible meaning of each word, reflecting the semantic depth encoded by lexical tone in Chinese languages. Furthermore, nearly all languages, whether tonal or non-tonal, employ contrastive pitch at the suprasegmental level (i.e., phrase or sentence) to express paralinguistic information (e.g., emotion, attitudes), predominantly through intonation, which is a principal component of prosody.^5^ This complexity showcases the pivotal and versatile role that pitch variation plays in the language comprehension and expression across different languages.

Other pitch variation features, such as lexical stress or accent (at the single word level) and prosody (at the phrase and sentences), have demonstrated significant diagnostic value in aphasia across languages.^6–11^ In non-tonal languages, lexical stress can differentiate meaning by emphasizing a particular syllable in multisyllabic words, typically through increased intensity, lengthened vowel duration, and changes in pitch.^12^ For instance, in English, the noun-verb distinction in “present” (/ˈprɛzᵊnt/ as a noun versus /prɛˈzᵊnt/ as a verb) hinges on syllabic/lexical stress placement. In languages where lexical stress conveys specific meaning, degradation of the semantic memory system may result in stress assignment errors, as observed in semantic variant (sv) primary progressive aphasia (PPA) in Italian and German speakers and among acquired aphasia of Russian speakers.^6–8^ Similarly, the literature on prosody has documented deterioration in both perception and production of prosody among English-speaking PPA patients.^9–11^ While there have been studies delineating the impairment in lexical stress and prosody intonation in PPA patients who are speakers of Indo-European languages, to the best of our knowledge, there have been no studies investigating lexical tone performance in PPA patients who speak tonal languages.

Laryngeal muscles, specifically the cricothyroid and thyroarytenoid muscles, regulate the length and tension of the vocal cords, which in turn control the amount of air passing through the vocal cords, thereby modulate the passive vibration of vocal cords and the pitch level of voice. For tonal languages, precise control of these muscles within the timescale of a single syllable is essential for generating a wide range of pitch variations.^13–15^ Therefore, it is unsurprising that neuroimaging and neurophysiological studies indicated the involvement of bilateral laryngeal motor cortex in the production of lexical tone.^16^ The bilateral superior temporal gyri has consistently been reported to play a key role in tone perception.^17–22^ Additionally, the involvement of bilateral anterior temporal, left inferior frontal gyri and precuneus has been noted, albeit less consistently reported.^17–22^ Many of these regions overlap with those affected in the three PPA variants. However, despite this anatomical convergence, our understanding of how such syndromes disrupt lexical tone processing remains limited. Specifically, the neural mechanisms underlying tone-related impairments and their potential diagnostic utility in clinical assessment warrant further investigation.

In this study, we aim to elucidate the patterns and neural underpinnings of lexical tone processing deficits in Mandarin- and Cantonese-speaking PPA patients. Given that tonal languages require precise temporal and articulatory control of the vocal folds and laryngeal muscles for accurate tone production, we hypothesize that Chinese-speaking individuals with the nonfluent/agrammatic variant PPA (nfvPPA) will exhibit impairments in lexical tone production in relation to motor planning deficits, apraxia of speech and fronto-insular atrophy typical of this syndrome in English speakers. Conversely, we speculate that Chinese-speaking logopenic variant (lv) PPA patients will produce more syllabic than tonal errors and may exhibit tone perception difficulties associated with phonological processing deficits and atrophy in the superior and posterior temporal regions. In contrast, individuals with semantic svPPA are expected to show impairments in the comprehension of lexical tones, corresponding to the characteristic anterior temporal lobe atrophy that disrupts semantic processing.

## Materials and methods

### Participants

This study included 78 native Chinese speakers from the international Chinese Language Assessment in PPA (CLAP) project, all of whom completed both motor speech and lexical tone assessments. Participants were recruited from seven sites in Hong Kong, Taiwan, and the United States. Of these, 24 were cognitively normal, the remaining 54 met current diagnostic criteria for PPA with 13 svPPA, 15 nfvPPA, 17 lvPPA and 9 PPA unclassifiable.^23^ Supplementary Figure 2 illustrates the atrophy patterns associated with each PPA variant. Participants either completed six years or more formal education in the Chinese languages or all their formal education were in Chinese if they had less than six years of formal education. Participants were excluded if they had other neurodegenerative diseases, a history of brain surgery, major brain trauma requiring hospitalization, oropharyngeal disorders affecting articulation, severe visual or hearing impairments, or a Mini-Mental State Exam score (MMSE)^24^ lower than 10. The study protocol was approved by the respective institutional review boards.

### Neuropsycholinguistic assessment

We collected MMSE^24^, CLAP battery and brain MRI imaging data from enrolled participants from all recruitment sites. As detailed in previous studies^25,26^, the CLAP battery consists of various speech and language tests, including confrontation naming, single-word comprehension, semantic association, syntax comprehension, repetition, and motor speech tests. In the CLAP confrontational naming test, participants were tasked with naming 48 pictures in which the items have high concreteness but varying word frequency. For the single-word comprehension test, participants were auditorily provided the names of 15 objects or animals and then asked to identify the target stimuli from a selection of six images, all belonging to the same semantic category. For the CLAP semantic association tests, participants were shown 30 sets of three words or pictures, with the target stimulus positioned above the other two stimuli. Participants were then asked to identify the words or pictures that best matched the target stimulus. In the syntax comprehension test, participants were presented with 30 Chinese sentences accompanied by corresponding pictures that varied in syntactic constructional meanings. Participants were then asked to select the picture that aligns with the semantic and syntactical meaning of each sentence. In the repetition task, participants were tasked with repeating both sensical and nonsensical phrases and sentences, ranging from three to eleven Chinese words in length. Assessments were conducted in each participant’s dominant language (Cantonese or Mandarin), as determined by language proficiency and usage. All assessments were administered by board-certified neurologists, neuropsychologists, speech-language pathologists, or supervised research staff, all of whom are native Mandarin or Cantonese speakers with over 12 years of formal education in the Chinese languages.

### Motor speech assessment

Studies of English-speaking individuals with nfvPPA have shown that apraxia of speech is particularly pronounced during the repetition of multisyllabic words that contain syllables with diverse places and manners of articulation and are rich in consonant clusters.^27^ The monosyllabic structure of Chinese characters and the relative absence of consonant clusters necessitate a tailored approach for eliciting motor speech impairments in Chinese speakers. To this end, we developed a motor speech battery in which participants are asked to repeat five distinct sets of phrases—each comprising four-or six-characters—five times consecutively. These phrases were systematically designed to assess specific articulatory features: (1) Place of articulation: phrases that varied in articulatory placement while holding tone and manner constant (e.g., Mandarin: 販售現畫 /fan4 shou4 xian4 hua4/; Cantonese: 答架八駕 /daap3 gaa3 baat3 gaa3/); (2) Manner of articulation: phrases that differed in how sounds are produced, with consistent articulatory placement and tone across syllables (e.g., Mandarin: 觀光開花 /guan1 guang1 kai1 hua1/; Cantonese: 花爸媽趴 /faa1 baa1 maa1 paa1/); (3) Tone of articulation: phrases that presented tonal contrasts across otherwise syllabically similar words (e.g., Mandarin: 麻媽罵馬 /ma2 ma1 ma4 ma3/; Cantonese: 引孕忍印 /jan5 jan6 jan2 jan3/), and (4) sensical phrases: idiomatic phrases that contrast in place, manner and tone of articulations (e.g., Mandarin: 防彈背心 /fang2 dan4 bei4 xin1/, Cantonese: 莫名奇妙 /mok6 ming4 kei4 miu6/), and (5) nonsensical phrases: phrases without lexical meaning constructed to contrast in place, manner, and tone for Cantonese speakers solely (e.g. Cantonese: 泰銀及寒 /taai3 ngan2 kap6 hon4). Notably, the tone of articulation phrases were specifically designed to evaluate tone production ability alongside motor speech performance, providing contrast to other articulatory dimensions. In addition to assessing repetition accuracy, we quantified the number of tonal and syllabic errors, as well as their proportion relative to each participant’s total errors during the motor speech assessment. To ensure accuracy and consistency in error coding, all speech recordings were independently reviewed by two native speakers of Mandarin and Cantonese.

### Lexical tone production assessment

Mandarin language encompasses four distinct tones: a high-level tone (Tone 1) and three contour tones—rising (Tone 2), dipping (Tone 3), and falling (Tone 4). In contrast, Cantonese features six contrasting tones: high-level (Tone 1), high-rising (Tone 2), mid-level (Tone 3), falling (Tone 4), low-rising (Tone 5), and low-level (Tone 6). Additionally, in Cantonese, when level tones (Tones 1, 3, and 6) end with a stop consonant, they are sometimes referred as checked tones (Tones 7, 8, 9).^28^ (Supplement Figure 1)

In addition to the repetition of tone-of-articulation phrases, lexical tone production was further assessed using two categories of tone twisters (i.e., phrases solely consist of words with identical or near-identical syllabic structures but varying lexical tones): sensical and nonsensical tone twisters (see Supplementary Table 1 for the full list). In the sensical stimuli, participants were asked to repeat two meaningful tone twister phrases in Mandarin or Cantonese, presented with both auditory and visual input. These stimuli, ranging from 8 to 12 words in length, consist of semantically coherent phrases with words that have similar syllables but differ in tones. Examples include the Mandarin one-syllable poem “石 室 詩 士 施 氏, 嗜 獅,誓 食 十 獅” /shi2 shi4 shi1 shi4 shi1 shi4, shi4 shi1, shi4 shi2 shi2 shi1/ and the Cantonese phrase “圓圓遠遠叫圓月” /yun4 yun4 yun5 yun5 giu3 yun4 yut6/. For the nonsensical stimuli, participants were presented with a sequence of words in both auditory and visual formats, each sharing similar syllables but differing in tone, such as “達/da2/ 大/da4/ 打/da3/ 搭/da1/” for Mandarin and “施 /si1/ 氏 /si6/ 市 /si5/ 時 /si4/ 史 /si2 事 /si6/” for Cantonese. We also included syllabically varied monotone phrases, wherein participants repeated phrases composed of words that shared similar tones but varied in their syllabic pronunciations. For example “風 淹鄉村” which is pronounced as /feng1 yan1 xiang1cun1/ in Mandarin and /fung1 yim1 heung1chyun1/ in Cantonese.

### Lexical tone perception and comprehension

To examine lexical tone perception and comprehension abilities, we designed four tests: Tone Discrimination, Tone Identification, Tone Picture Matching, and Tone Word Matching tests. In the Tone Discrimination Test, participants were presented with pairs of single syllable sounds in audio format. These sounds either carried the same tones (e.g., ma2-ma2) or different tones (e.g., ma2-ma4), and participants were asked to judge whether the tones were the same or different. This task primarily engages tone perceptual processing, involving same–different judgment of isolated tonal contrasts in the absence of semantic context. The Tone Identification Test constituted a unimodal semantic recognition task by presenting participants with audio stimuli consisting of single syllable sounds carrying tones from Mandarin, Cantonese, Thai, and Vietnamese languages. Mandarin speakers were asked to determine if the twelve sounds presented were Mandarin tones, while Cantonese-speaking participants were asked to identify twenty sounds in Cantonese tones. Cantonese speakers were given more stimuli due to the greater number of tones in the language. Finally, the Tone Picture Matching and Tone Word Matching Tests were designed as transmodal semantic comprehension measures. Participants were presented with single syllable sounds and four pictures or words that shared the same syllable sounds but varied in tones (e.g., 兔 /tu4/土 /tu2/ 禿 /tu1/ 圖 /tu3/) and then asked to match the sounds with the corresponding pictures or words. These tasks rely heavily on lexical-semantic processing, requiring integration of tonal and semantic information for accurate comprehension.

### Statistical Analysis

Demographic features were compared across diagnostic groups using analysis of variance (ANOVA) for continuous variables, with Bonferroni correction for multiple comparisons, and Pearson’s Chi-squared tests for categorical variables. Group differences in MMSE and CLAP battery performance, including error patterns, were assessed using nonparametric Kruskal–Wallis H tests, given that neuropsychological data did not meet the assumptions of normality and homogeneity of variance, as indicated by Shapiro–Wilk tests. For tests that yielded statistically significant diagnostic group effects (*p* < 0.05), pairwise post-hoc comparisons were performed with Bonferroni adjustment to control for family-wise error. To further account for confounding effects of age, education, and testing language (Cantonese vs. Mandarin), we applied Generalized Estimating Equations (GEE) with a binomial distribution and logit link function to analyze lexical tone and motor speech scores. The dependent variable was defined as the proportion of correct responses per task, allowing for variation in trial counts across language groups. Diagnosis and testing language were entered as fixed effects, while age and years of education were included as covariates. Robust standard errors were employed, and subject IDs were specified to ensure accurate variance estimation. Type III Wald chi-square tests were used to evaluate the significance of model predictors. Estimated marginal means (EMMs) were computed for diagnostic groups and testing language, adjusted for age and education. Pairwise comparisons between EMMs were performed with Bonferroni correction for multiple comparisons. Model fit was evaluated using the Quasi-likelihood under the Independence Model Criterion and its corrected variant.

To explore the diagnostic value of lexical tone performance in PPA, we additionally examined lexical tone measures using nonparametric ROC analyses. Specifically, tone production and tonal errors were evaluated for identifying nfvPPA, tone comprehension for distinguishing svPPA, and syllabic errors for detecting lvPPA, as hypothesized. For each task, we computed the area under the curve (AUC), sensitivity, specificity, and optimal cutoff points using Youden’s Index. Furthermore, a nonparametric Spearman’s rank-order correlation was used to examine the correlation between the three lexical tone and syllable production tasks. All statistical analyses were conducted using IBM SPSS Statistics, version 27.0 (IBM Corp.), with a two-tailed significance threshold set at α = 0.05.

### Brain behavioral correlation

To explore the neural mechanism underlying lexical tone performances, we examined the neural correlates in participants who completed the CLAP battery and MRI within a three-month interval. This comprised 10 participants with svPPA, 11 participants with nfvPPA, 12 patients with lvPPA, 7 patients with unclassifiable PPA, and 15 cognitively normal controls. MRI sequences were acquired with 3T scanners following the Alzheimer’s Disease Neuroimaging Initiative 3 protocol (ADNI 3; http://adni.loni.usc.edu/adni-3/). The ADNI 3 imaging protocol included a high-resolution T1-weighted 3D magnetization prepared rapid acquisition gradient echo structural scan with the following parameters: 200-211 sagittal slices, repetition time =2300 ms, echo time =2.98 ms, inversion time = 900 ms, flip angle 9°–11°, field of view = 256 mm^3^, matrix size = 256×256, in-plane voxel size 1.0 × 1.0 mm^2^; slice thickness = 1 mm. T1-weighted images were preprocessed using Computational Anatomy Toolbox (CAT12) in the Statistical Parametric Mapping software (SPM), operating under Matlab 2022b. Preprocessing steps included bias-field correction, skull stripping, and tissue segmentation into gray matter (GM), white matter, and cerebrospinal fluid, using an adaptive maximum a posteriori algorithm. GM probability maps were nonlinearly normalized to the Montreal Neurologic Institute space using DARTEL, modulated via the Jacobian determinant from the spatial normalization, and smoothed by an isotropic Gaussian kernel of 8 mm full-width at half-maximum.

To examine the neural correlates of lexical tone performance, we conducted voxel-based morphometry (VBM) analysis using the Statistical NonParametric Mapping (SnPM13) toolbox implemented in SPM12 and MATLAB^29,30^. A regression model was employed to assess the association between gray matter volume and the performance of lexical tone tests, with individual performance scores as the primary covariate and age at examination, sex, total gray matter volume, years of education, and testing language (Mandarin or Cantonese) included as covariates. Nonparametric statistical inference was performed with 5,000 permutations. A cluster-wise inference approach was adopted, applying a cluster-forming threshold of t = 3.09 (uncorrected p < 0.001). Family-wise error (FWE) correction was performed at the cluster level with a corrected alpha of 0.05^31^.

### Data availability

Anonymized data are available from the corresponding author upon request and subject to approval and completion of a Material Transfer Agreement. Data are not publicly accessible in accordance with institutional ethical protocols governing participant confidentiality.

## Results

### Demographic and neurolinguistic profiles

Table 1 summarized the demographic and linguistic profiles across the study groups. Demographically, no significant differences were observed in handedness *[χ²(8,78)= 8.82, p = 0.357*] or sex distribution [*χ²(4,78)= 9.34, p = 0.053*], although age at examination differed between control and PPA unclassifiable groups [*F(4,73)= 2.74, p* = 0.035]. Years of education also varied significantly among groups [*F(4,73)= 5.71, p < 0.001*], with the control group attaining significantly more years of formal education compared to individuals with lvPPA and PPA unclassifiable.

**Table 1.** Demographic characteristics and neuropsychological performances of the study participants (n=78).

|  | Control<br>(n= 24) | svPPA<br>(n= 13) | nvPPA<br>(n= 15) | lvPPA<br>(n= 17) | Unclassifiable PPA<br>(n= 9) | p-value |
| --- | --- | --- | --- | --- | --- | --- |
| **Demographic characteristics** |  |  |  |  |  |  |
| Age at examination (years) | 65.88 (7.10)<sup>d</sup> | 68.08 (5.36) | 67.33 (6.73) | 69.47 (7.41) | 74.00 (3.9)<sup>d</sup> | 0.035 |
| Sex (Female/ Male) | 15/9 | 5/8 | 11/4 | 5/12 | 3/6 | 0.053 |
| Years of education | 16.67 (3.62)<sup>c,d</sup> | 14.12 (5.00) | 13.73 (3.22) | 10.94 (3.89)<sup>c</sup> | 12.00(4.74)<sup>d</sup> | <0.001 |
| Handedness (Right/Left/Ambidextrous) | 22/0/2 | 13/0/0 | 14/1/0 | 17/0/0 | 9/0/0 | 0.357 |
| **Global Cognition** |  |  |  |  |  |  |
| Mini-Mental State Exam (MMSE)<sup>k</sup> | 30 (1)<sup>a,b,c,d</sup> | 21 (8)<sup>a</sup> | 27 (5)<sup>b</sup> | 20 (7)<sup>c</sup> | 25 (7)<sup>d</sup> | <0.001 |
| **Speech and Language** |  |  |  |  |  |  |
| Picture Naming (0-48) | 48 (1)<sup>a,c</sup> | 4.50 (8)<sup>a,e,g</sup> | 45.50 (16)<sup>e</sup> | 32.50 (27)<sup>c</sup> | 42 (13)<sup>g</sup> | <0.001 |
| Single Word Comprehension (0-15)<sup>l</sup> | 15 (1)<sup>a,b,c</sup> | 6.50 (5)<sup>a,e,g</sup> | 14 (2)<sup>b,e</sup> | 13 (3)<sup>c</sup> | 14 (2)<sup>g</sup> | <0.001 |
| Semantic Association: Picture (0-15)<sup>m</sup> | 15 (0)<sup>a,c</sup> | 9.50 (3)<sup>a,e,g</sup> | 14.50 (2)<sup>e</sup> | 13 (3)<sup>c</sup> | 14.50 (1)<sup>g</sup> | <0.001 |
| Semantic Association: Word (0-15)<sup>n</sup> | 15 (0)<sup>a,c</sup> | 9.50 (5)<sup>a,e,g</sup> | 14.50 (2)<sup>e</sup> | 13 (3)<sup>c</sup> | 15 (1)<sup>g</sup> | <0.001 |
| Syntax Comprehension (0-30)<sup>o</sup> | 30 (0)<sup>a,b,c</sup> | 27.5 (4)<sup>a</sup> | 26.0 (3)<sup>b</sup> | 22.5 (5)<sup>c</sup> | 29 (5)<sup>d</sup> | 0.001 |
| Sentence Repetition (0-100)<sup>l</sup> | 94.0 (6.8)<sup>a,b,c,d</sup> | 79.7 (13.3)<sup>a</sup> | 84.4 (8.3)<sup>b</sup> | 66.2 (27.5)<sup>c</sup> | 80.6 (21.2)<sup>d</sup> | <0.001 |

<sup>a</sup> Significant between Control and svPPA; <sup>b</sup> Significant between Control and nvPPA; <sup>c</sup> Significant between Control and lvPPA; <sup>d</sup> Significant between control and unclassifiable PPA; <sup>e</sup> Significant between svPPA and nvPPA; <sup>f</sup> Significant between svPPA and lvPPA; <sup>g</sup> Significant between svPPA and unclassifiable PPA; <sup>h</sup> Significant between nvPPA and lvPPA; <sup>i</sup> Significant between nvPPA and unclassifiable PPA; <sup>j</sup> Significant between unclassifiable PPA and lvPPA; <sup>k</sup> Control (n=23), svPPA (n=13), nvPPA (n=15), unclassifiable PPA (n=9), lvPPA (n=17); <sup>l</sup> Control (n=23), svPPA (n=13), nvPPA (n=14), unclassifiable PPA (n=9), lvPPA (n=17); <sup>m</sup> Control (n=24), svPPA (n=11), nvPPA (n=14), unclassifiable PPA (n=9), lvPPA (n=17); <sup>n</sup> Control (n=23), svPPA (n=11), nvPPA (n=14), unclassifiable PPA (n=9), lvPPA (n=17); <sup>o</sup> Control (n=22), svPPA (n=11), nvPPA (n=13), unclassifiable PPA (n=9), lvPPA (n=17)

In terms of global cognition, as measured by the Mini-Mental State Examination (MMSE), all PPA subtypes showed significantly lower scores relative to controls [*H(4)= 54.24, p < 0.001*]. Among the speech and language tests, patients with svPPA exhibited marked impairment in tasks dependent on semantic knowledge, specifically picture naming [*H(4)= 55.42, p < 0.001*], single-word comprehension [*H(4)= 48.13, p < 0.001*], and both semantic association tasks [Picture: *H(4)=43.91, p < 0.001*; Word: *H(4)=39.88, p < 0.001*]. Patients with lvPPA also performed significantly worse than controls on semantic tests, though their scores remained significantly higher than those of the svPPA group. Syntax comprehension and sentence repetition was notably reduced in the PPA groups compared to controls [Syntax: *H(4)=34.65, p < 0.001*; Repetition: *H(4)=40.81, p < 0.001*], with the lvPPA group exhibited lowest performance.

### Motor Speech performance

Figure 1 and Table 2 illustrated the performance of each diagnostic group on tasks assessing lexical tone production, lexical tone perception, and motor speech functions.

**Figure 1.**
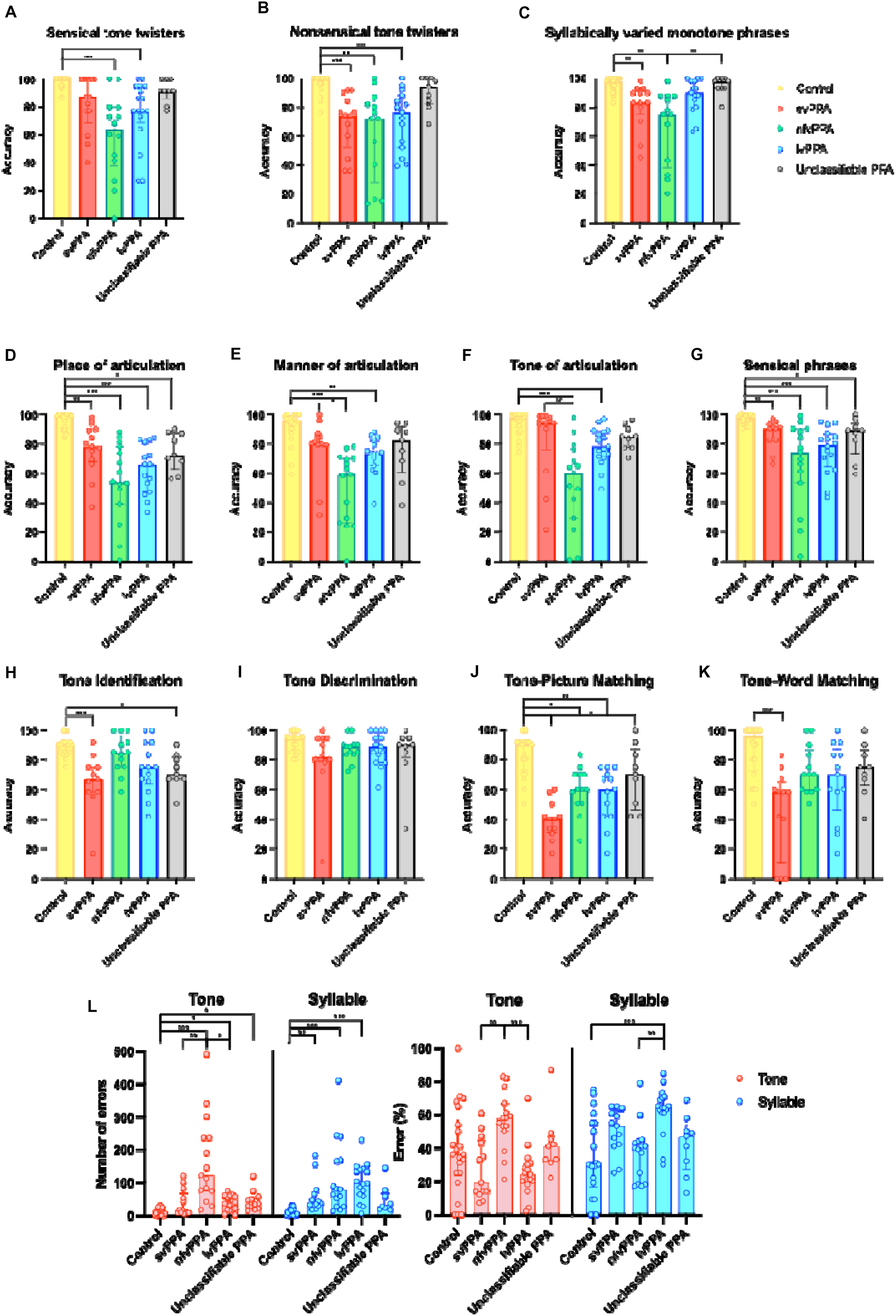
Lexical tone and motor speech performance across diagnostic groups. Presented are the results of lexical tone and motor speech performance tasks. Tone and syllable production tasks include: **(A)** Sensical tone twisters; **(B)** Nonsensical tone twisters; **(C)** Syllabically varied monotone phrases. Motor speech assessment comprises: **(D)** Place of articulation; **(E)** Manner of articulation; **(F)** Tone of articulation; **(G)** Sensical phrases. Tone perception is evaluated using: **(H)** Tone identification; **(I)** Tone discrimination; **(J)** Tone– picture matching; **(K)** Tone–word matching. **(L)** shows the frequency and proportional distribution of tone and syllable errors. Bars represent group medians and interquartile ranges; individual participant data are overlaid as scatter plots. Statistical comparisons were performed using the Kruskal–Wallis test with Bonferroni-adjusted post hoc comparisons (*p* < 0.05; p < 0.01; *p* < 0.001).

**Table 2.**
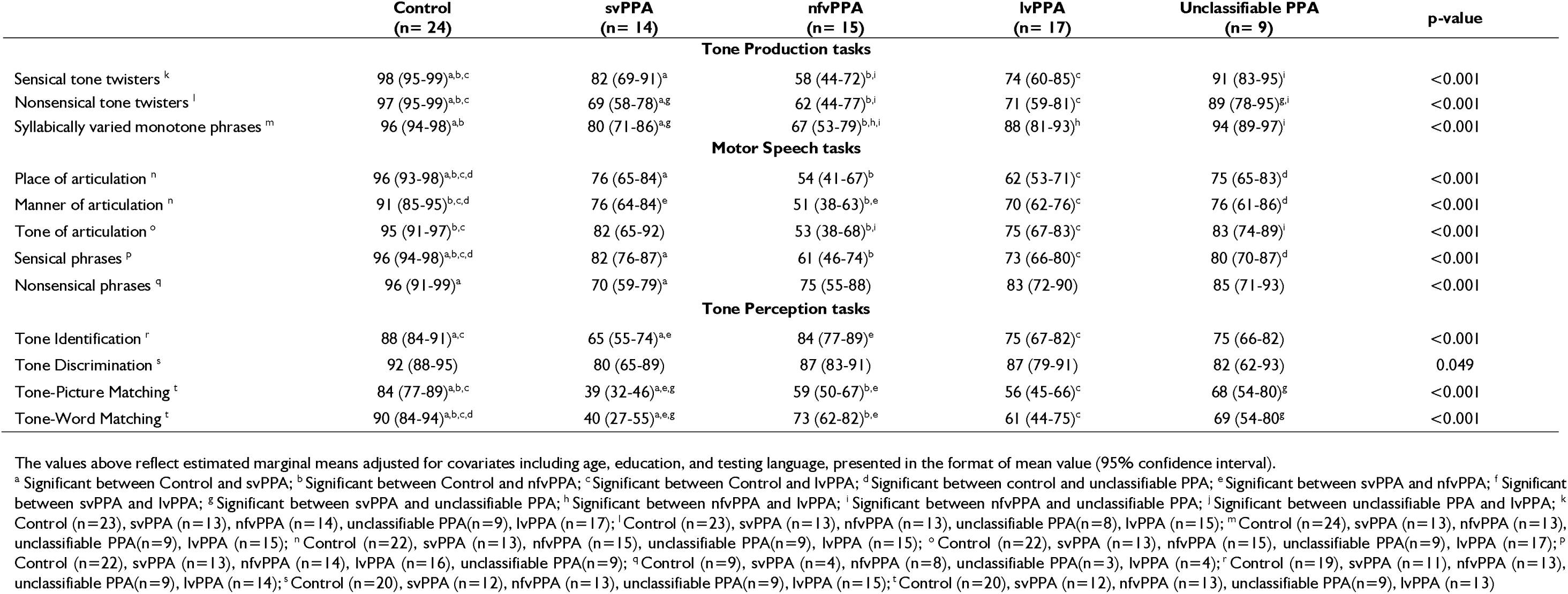
The performance of the CLAP lexical tones tests among the study participants adjusted for age, education and testing languages (n=78).

In the motor speech assessment, significant diagnostic group differences were observed in the repetition of phrases varying in place, manner, and tone of articulation, as well as in sensical and nonsensical idiomatic expressions [Place: *H (4) = 45.04, p < 0.001*; Manner: *H (4) = 36.52, p < 0.001*; Tone: *H (4) = 34.41, p < 0.001*; Sensical: *H (4) = 37.20, p < 0.001;* Nonsensical: *H (4) = 13.76, p = 0.008*]. Specifically, all PPA variants exhibited significantly reduced accuracy relative to controls in repeating phrases that differed in place of articulation before and after adjusting for demographic and language-related covariates [Wald *χ²(4) = 61.35, p < .001*].

When producing phrases that contrasted in manner and tone of articulation, both nfvPPA and lvPPA participants demonstrated significantly lower accuracy compared to controls, with nfvPPA individuals additionally performing worse than those with svPPA. After adjusting for age, education, and testing language, diagnostic group differences remained significant for manner articulation tasks [*Wald χ²(4) = 35.25, p < .001*]. Participants with nfvPPA showed the most pronounced impairments, with a 41% reduction in accuracy compared to controls (*p* < .001). Significant differences were also observed between controls and lvPPA (−21%, p < .001), controls and svPPA (−16%, p = .047), and between nfvPPA and svPPA (−25%, p = .037). For phrases differing in tone, diagnostic group differences also remained significant following covariate adjustment [Wald χ²(4) = 40.35, *p* < .001]. Pairwise comparisons revealed that nfvPPA participants performed significantly worse than controls (*−41%, p < 0.001*) and unclassifiable PPA (*−30%, p = 0.004*), with marginal differences relative to svPPA (*−29%, p = 0.060*) and lvPPA (*−30%, p =0 .081*).

When asked to produce sensical idiomatic expressions, controls significantly outperformed all PPA groups. After adjusting for covariates, the effect of diagnosis remained significant [*Wald χ²(4) = 51.48*]. Relative to controls, significant accuracy reductions were observed in lvPPA *(−23%, p < 0.001*), nfvPPA (*−35%, p < 0.001*), svPPA (*−14%, p < 0.001*), and unclassifiable PPA (*−16%, p = 0.003*). For the repetition of nonsensical phrases, although diagnostic group differences were observed, none of the post-hoc pairwise comparisons reached statistical significance. After adjusting for covariates, controls significantly outperformed the unclassifiable PPA group (*-26%, p < 0.001*) while other between-group differences did not reach significance after Bonferroni correction. Interestingly, while clinical diagnosis remained a robust predictor of performance across all these phrase types, testing language emerged as a significant predictor only for tone of articulation phrases [*Wald χ²(1) = 4.93, p = 0.026*], with Mandarin speakers outperforming Cantonese speakers (Mandarin: *86%, 95% CI: 81–89%*; Cantonese: *75%, 95% CI: 65–83%; p = .041*).

ROC analyses revealed that that production of phrases varying in manner of articulation achieved strong classification for nfvPPA (*AUC = 0.882; sensitivity=100%, specificity = 67.8%, cutoff = 78.5%*). Tone of articulation phrases also showed robust discrimination (*AUC = 0.860; sensitivity = 93.3%, specificity = 72.9%, cutoff = 68.6%*), while place of articulation phrase yielded a moderate AUC of 0.793 (*sensitivity = 86.7%, specificity = 57.6%, cutoff = 81.5%*). In contrast, repetition of sensical phrases demonstrated weaker classification accuracy (*AUC = 0.702; sensitivity = 93.3%, specificity = 49.2%*).

Analyses of error types in the motor speech assessement further revealed pronounced group differences across PPA variants and controls [*H(4) = 38.58, p < 0.001*]. Tonal error counts were significantly lower in controls relative to all PPA subtypes, with the exception of svPPA. Additionally, patients with nfvPPA produced the highest number of tonal errors, significantly exceeding those with control (*p < 0.001*), svPPA (*p = 0.007*) and lvPPA (*p = 0.04*). With respect to syllabic errors, svPPA, nfvPPA and lvPPA patients exhibited significantly more errors than control participants [*H(4) =41.06, P < 0.001*]. When examining the proportional distribution of error types, nfvPPA also demonstrated a significantly higher proportion of tonal error relative to svPPA and lvPPA groups [*H(4) = 20.64, P < 0.001*]. In contrast, lvPPA was characterized by disproportionately elevated syllabic error percentages relative to control and nfvPPA groups [*H(4) = 22.13, P < 0.001*]. These findings suggest that while individuals with nfvPPA and lvPPA produced substantial numbers of tonal and syllabic errors, tonal errors were disproportionately more frequent in nfvPPA, whereas syllabic errors predominated in lvPPA.

ROC analyses revealed that tone error count was a strong discriminator of nfvPPA from other diagnostic groups, yielding an AUC of 0.907 (*sensitivity = 91.8%, specificity = 80% at a cutoff of ≥68.5 errors*). Tone error percentage also showed good discriminative ability (*AUC = 0.810; sensitivity = 82%, specificity = 80% at ≥49.5%*). For lvPPA, syllable error count distinguished it from other groups with an AUC of 0.771 (*sensitivity = 82.4%, specificity = 72.9% at ≥58.5 errors*), with syllable error percentage yielding similar discriminative profile (*AUC = 0.834; sensitivity = 82.4%, specificity = 84.7% at ≥60.75%*).

### Lexical tone production performance

In reciting sensical tone twisters, significant differences were observed across the diagnostic groups [*H(4)= 29.38, p < 0.001*], with individuals in the nfvPPA and lvPPA groups demonstrating significantly lower accuracy compared to controls (both *p ≤ 0.001*). This diagnostic group effect remained robust after adjusting for age, education, and testing language, with diagnosis remained a significant predictor of sensical tone twister scores [*Wald χ²(4) = 54.35, p < .001*]; in contrast, testing language, age, and education were not significant covariates. Estimated marginal means (EMM), adjusted for the covariates, revealed that the nfvPPA group exhibited the lowest adjusted accuracy (Table 2). Pairwise comparisons further indicated that individuals with nfvPPA performed significantly worse than controls (mean difference = –39%, p < .001) and those with unclassifiable PPA (–32%, p = .001). The difference between nfvPPA and svPPA was marginal *(-24%, p = .065*), while the contrast with lvPPA was non-significant.

A similar pattern was observed in the producing nonsensical tone twisters, where controls significantly outperformed the svPPA, nfvPPA, and lvPPA groups [*H(4) = 35.30, p < 0.0010*]. After accounting for demographic and language effects, diagnosis remained a significant predictor (*Wald χ²(4) = 55.03, p < .001*), whereas testing language, age, and education were not statistically significant. Adjusted EMMs revealed the highest accuracy among controls, followed by unclassifiable PPA, svPPA, lvPPA, and the lowest in nfvPPA. Pairwise comparisons showed that controls also significantly outperformed svPPA (mean difference = –28%, p < .001), lvPPA (–26%, p < .001), and nfvPPA (–36%, p < .001).

A comparable trend was also evident in the production of syllabically varied monotone phrases [*H(4)= 33.07, p < 0.001*], with controls achieving higher accuracy than both svPPA and nfvPPA participants (both *p ≤ 0.001*), and the nfvPPA group also demonstrating significantly lower scores than the unclassifiable group (*p = 0.005*). After adjusting for demographic and language variables, diagnosis remained a significant predictor. EMMs were highest in the control group, followed by unclassifiable PPA, lvPPA, svPPA, and nfvPPA. Post hoc pairwise comparisons indicated that nfvPPA participants performed significantly worse than all other groups other than svPPA (*all p ≤ 0.045*).

ROC analysis revealed that lexical tone production tasks demonstrated significant discriminatory ability in identifying individuals with nfvPPA. The sensical tone twister task yielded an AUC of 0.790 (*95% CI: 0.638–0.942*), with 84.6% sensitivity and 76.3% specificity at a cutoff of 80.91%. The nonsensical tone twister showed lower discriminative performance (*AUC = 0.724, 95% CI: 0.570–0.878*), with 76.9% sensitivity and 62.7% specificity at a cutoff of 83.03%. In contrast, the monotone syllabically varied phrase task achieved the highest classification accuracy (*AUC = 0.851, 95% CI: 0.744–0.959*), with 92.3% sensitivity and 72.1% specificity at an optimal cutoff of 88.75%. All three tone production tasks were significantly positively correlated: sensical and nonsensical tone twisters (*ρ = 0.664, p < .001*), nonsensical tone twisters and monotone syllabically varied phrases (*ρ = 0.756, p < .001*), and sensical tone twisters and monotone phrases (*ρ = 0.628, p < .001*).

### Lexical tone perception and comprehension performance

In the tone identification task, svPPA and unclassifiable PPA participants demonstrated significantly lower accuracy than controls [*H(4)= 20.21, p < 0.001*]. After covariates adjustment, the performance deficit in the svPPA group remained significant compared to control and nfvPPA [*Wald χ²(4) = 32.76, p < .001*]. Post-hoc comparisons revealed that individuals with svPPA performed significantly worse than controls (*−22%, p < .001*) and nfvPPA (*−19%, p = .018*). Additionally, lvPPA participants showed reduced accuracy compared to controls (*−13%, p = 0.031*). In contrast, no significant group differences were observed in the tone discrimination task (*p = 0.206*). Although diagnostic group remained a significant predictor after adjusting for covariates (*p = 0.049*), none of the pairwise comparisons reached statistical significance following correction.

In the tone–picture matching test, controls outperformed most PPA groups, with svPPA participants also scoring significantly lower than unclassifiable PPA groups [*H(4) = 34.14, p < 0.001*]. After adjusting for age, education, and testing language, diagnosis remained a significant predictor of tone–picture matching performance (*Wald χ²(4) = 62.77, p < 0.001*). Specifically, svPPA participants performed significantly worse than controls *(−46%, p < 0.001*), nfvPPA (*−20%, p = 0.006*), and lvPPA (*−30%, p = 0.001*). Controls also significantly outperformed lvPPA (*−28%, p < 0.001*) and unclassifiable PPA participants (*−25%, p < 0.001*). Similarly, in the tone–word matching test, individuals with svPPA showed significantly lower accuracy than controls [*H(4) = 19.41, p < 0.001*]. GEE revealed significant effects of diagnosis (*Wald χ²(4) = 39.11, p < 0.001*) and testing language (*Wald χ²(1) = 12.48, p < 0.001*) on tone-word matching accuracy. Notably, controls significantly outperformed all PPA variants, including svPPA (*-49%, p < 0.001*), nfvPPA (*-16%, p = 0.042*), lvPPA (*-29%, p = 0.005*), and unclassifiable PPA (*-21%, p = 0.035*). Additionally, svPPA participants performed significantly worse than both nfvPPA (*-33%, p = 0.005*) and unclassifiable PPA (*-28%, p = 0.023*). Regarding testing language, Mandarin-speaking participants outperformed Cantonese speakers (*78% vs. 58%, p < .001*).

ROC analysis showed that tone picture matching achieved the highest classification accuracy for svPPA (*AUC = 0.884; sensitivity = 100%, specificity = 63.0% at a 59.2% cutoff*). Tone word matching also performed well (*AUC = 0.809; sensitivity = 100%, specificity = 37.0%*). Tone identification yielded an AUC of 0.769 (*sensitivity = 90.9%, specificity = 48.1%*), while tone discrimination showed lower accuracy (*AUC = 0.624; sensitivity = 54.5%, specificity = 83.3% at an 81.7% cutoff*).

### Neuroanatomical correlation of lexical tone scores

The VBM results for lexical tone production and perception tests are summarized in Figure 2 and Table 3. In summary, nonsensical tone twister production was significantly associated with gray matter volume in the left superior and middle temporal, inferior frontal, medial temporal, fusiform gyri and insula. While neural correlate analyses of sensical tone twisters and syllabically varied monotone phrases did not survive FWE correction at the cluster level (*p < 0.05*), exploratory analyses at an uncorrected threshold (*p < 0.05*) revealed positive associations for sensical tone twisters in the left pars triangularis and insula, and for syllabically varied monotone phrases in the left pars opercularis, insula, and superior temporal gyrus. Upon consolidating the findings across the three tone and syllable production tasks, overlapping regions were consistently localized to the left insula (Figure 2D). Conversely, no significant gray matter associations were identified in the VBM analyses of the motor speech assessment tasks.

**Figure 2.**
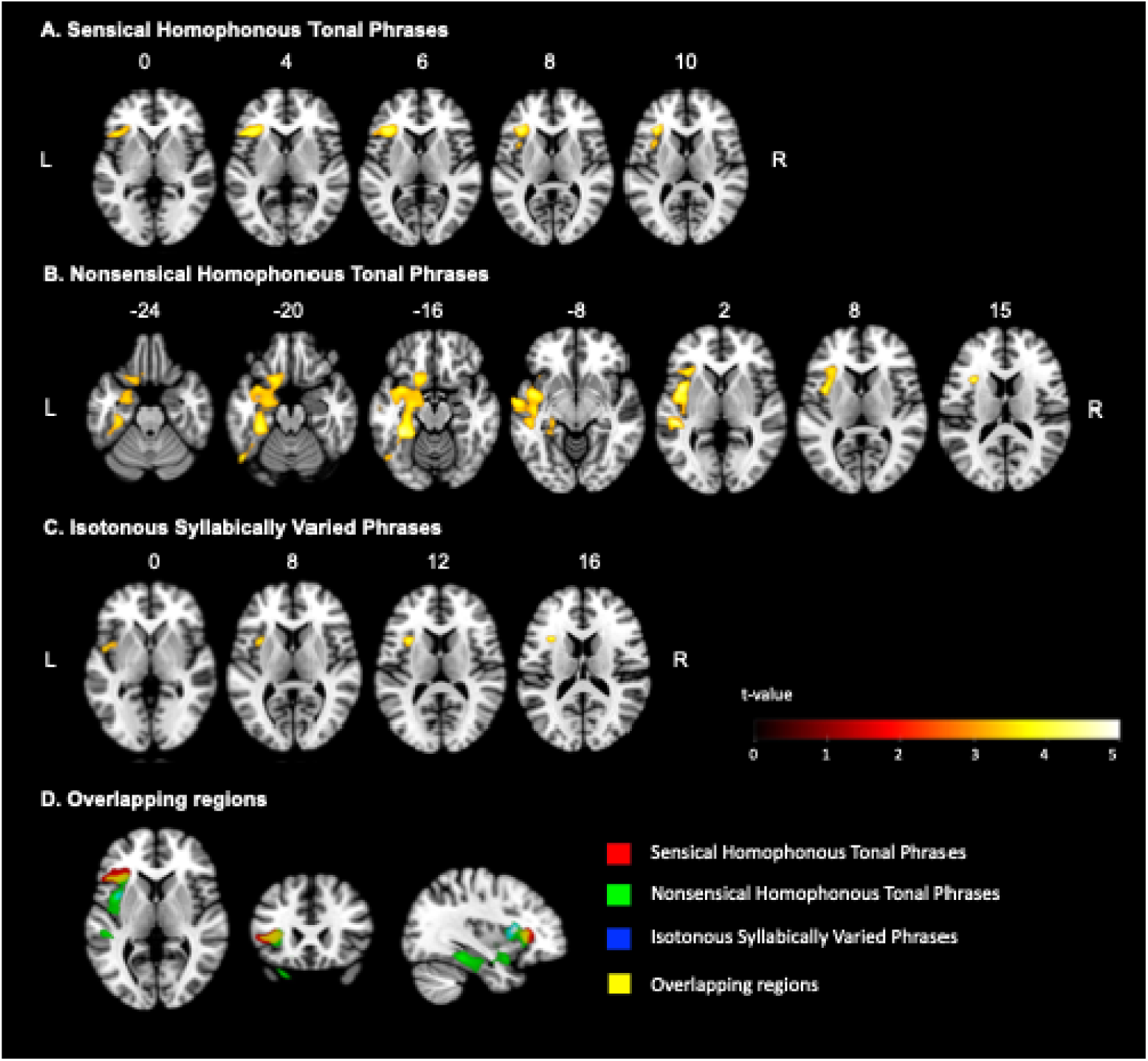
Neuroanatomical correlates of lexical tone and syllable production tests. The figure shows the voxel-based morphometry analyses of **(A)** Sensical homophonous tonal phrases; **(B)** Nonsensical homophonous tonal phrases; **(C)** Isotonous syllabically varied phrases; with **(D)** indicating overlapping regions across the three production tasks. For **(A)∼(C),** statistical parametric maps show brain regions where greater gray matter volume was significantly associated with higher task performance. Analyses were conducted using SnPM13, with age, sex, total gray matter volume, education, and testing language as covariates. Nonparametric inference (5,000 permutations) used a cluster-forming threshold of t = 3.09 (uncorrected p < 0.001), with family-wise error correction at the cluster level (α = 0.05) and an extent threshold of k > 100. Slices are labeled with MNI coordinates (L = left, R = right); the color bar represents t-values. For (D), red, green, and blue indicate task-specific associations, and yellow highlights overlapping regions.

**Figure 3.**
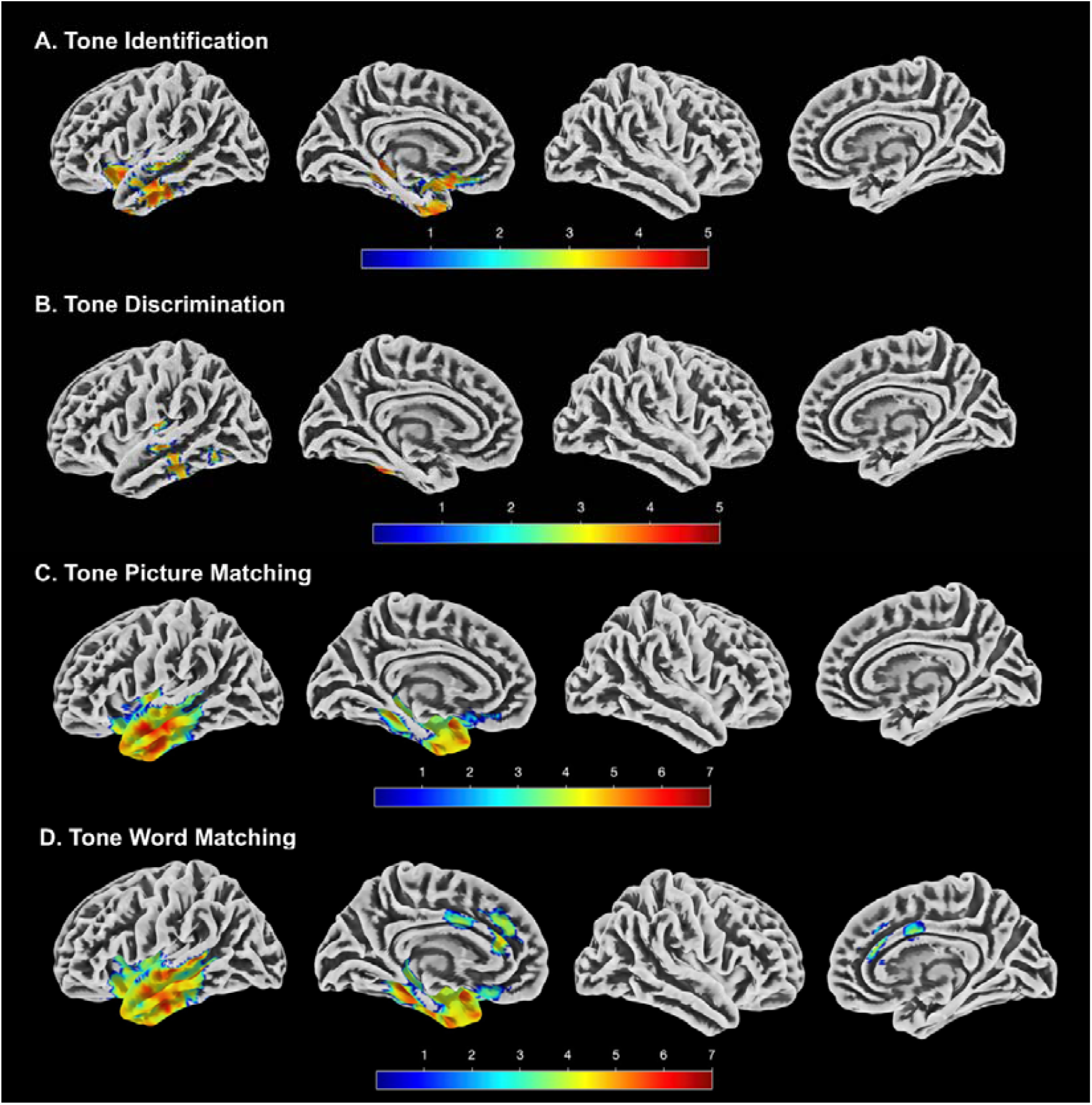
Neuroanatomical correlates of lexical tone perception tests. This brain maps display regions where gray matter volume was significantly associated with performance on: **(A)** Tone identification, **(B)** Tone discrimination, **(C)** Tone picture matching, and **(D)** Tone word matching. Voxel-based morphometry analyses were conducted using SnPM13 with age, sex, total gray matter volume, years of education, and testing language as covariates. Nonparametric inference employed 5,000 permutations with a cluster-forming threshold of t = 3.09 (uncorrected p < 0.001) and family-wise error correction at the cluster level (α = 0.05). The color bar represents t-values for each task.

**Table 3.**
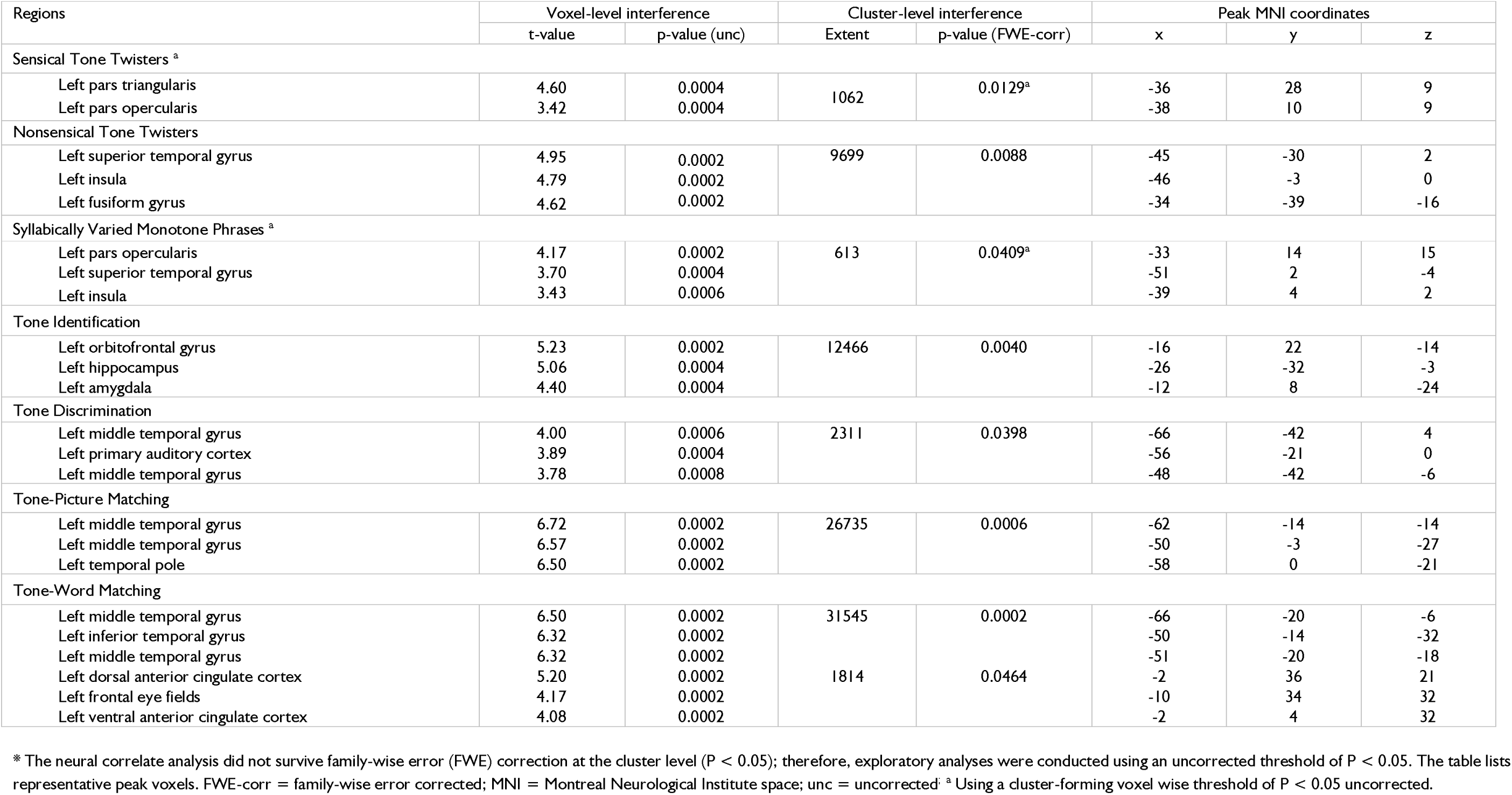
Neuroanatomical correlates of tone production and perception tests.

| Regions | Voxel-level interference |  | Cluster-level interference |  | Peak MNI coordinates |  |  |
| --- | --- | --- | --- | --- | --- | --- | --- |
|  | t-value | p-value (unc) | Extent | p-value (FWE-corr) | x | y | z |
| Sensical Tone Twisters <sup>a</sup> |  |  |  |  |  |  |  |
| Left pars triangularis | 4.60 | 0.0004 | 1062 | 0.0129<sup>a</sup> | -36 | 28 | 9 |
| Left pars opercularis | 3.42 | 0.0004 |  |  | -38 | 10 | 9 |
| Nonsensical Tone Twisters |  |  |  |  |  |  |  |
| Left superior temporal gyrus | 4.95 | 0.0002 | 9699 | 0.0088 | -45 | -30 | 2 |
| Left insula | 4.79 | 0.0002 |  |  | -46 | -3 | 0 |
| Left fusiform gyrus | 4.62 | 0.0002 |  |  | -34 | -39 | -16 |
| Syllabically Varied Monotone Phrases <sup>a</sup> |  |  |  |  |  |  |  |
| Left pars opercularis | 4.17 | 0.0002 | 613 | 0.0409<sup>a</sup> | -33 | 14 | 15 |
| Left superior temporal gyrus | 3.70 | 0.0004 |  |  | -51 | 2 | -4 |
| Left insula | 3.43 | 0.0006 |  |  | -39 | 4 | 2 |
| Tone Identification |  |  |  |  |  |  |  |
| Left orbitofrontal gyrus | 5.23 | 0.0002 | 12466 | 0.0040 | -16 | 22 | -14 |
| Left hippocampus | 5.06 | 0.0004 |  |  | -26 | -32 | -3 |
| Left amygdala | 4.40 | 0.0004 |  |  | -12 | 8 | -24 |
| Tone Discrimination |  |  |  |  |  |  |  |
| Left middle temporal gyrus | 4.00 | 0.0006 | 2311 | 0.0398 | -66 | -42 | 4 |
| Left primary auditory cortex | 3.89 | 0.0004 |  |  | -56 | -21 | 0 |
| Left middle temporal gyrus | 3.78 | 0.0008 |  |  | -48 | -42 | -6 |
| Tone-Picture Matching |  |  |  |  |  |  |  |
| Left middle temporal gyrus | 6.72 | 0.0002 | 26735 | 0.0006 | -62 | -14 | -14 |
| Left middle temporal gyrus | 6.57 | 0.0002 |  |  | -50 | -3 | -27 |
| Left temporal pole | 6.50 | 0.0002 |  |  | -58 | 0 | -21 |
| Tone-Word Matching |  |  |  |  |  |  |  |
| Left middle temporal gyrus | 6.50 | 0.0002 | 31545 | 0.0002 | -66 | -20 | -6 |
| Left inferior temporal gyrus | 6.32 | 0.0002 |  |  | -50 | -14 | -32 |
| Left middle temporal gyrus | 6.32 | 0.0002 |  |  | -51 | -20 | -18 |
| Left dorsal anterior cingulate cortex | 5.20 | 0.0002 | 1814 | 0.0464 | -2 | 36 | 21 |
| Left frontal eye fields | 4.17 | 0.0002 |  |  | -10 | 34 | 32 |
| Left ventral anterior cingulate cortex | 4.08 | 0.0002 |  |  | -2 | 4 | 32 |

\* The neural correlate analysis did not survive family-wise error (FWE) correction at the cluster level ( $P < 0.05$ ); therefore, exploratory analyses were conducted using an uncorrected threshold of $P < 0.05$ . The table lists representative peak voxels. FWE-corr = family-wise error corrected; MNI = Montreal Neurological Institute space; unc = uncorrected; <sup>a</sup> Using a cluster-forming voxel wise threshold of $P < 0.05$ uncorrected.

**Table 4.**
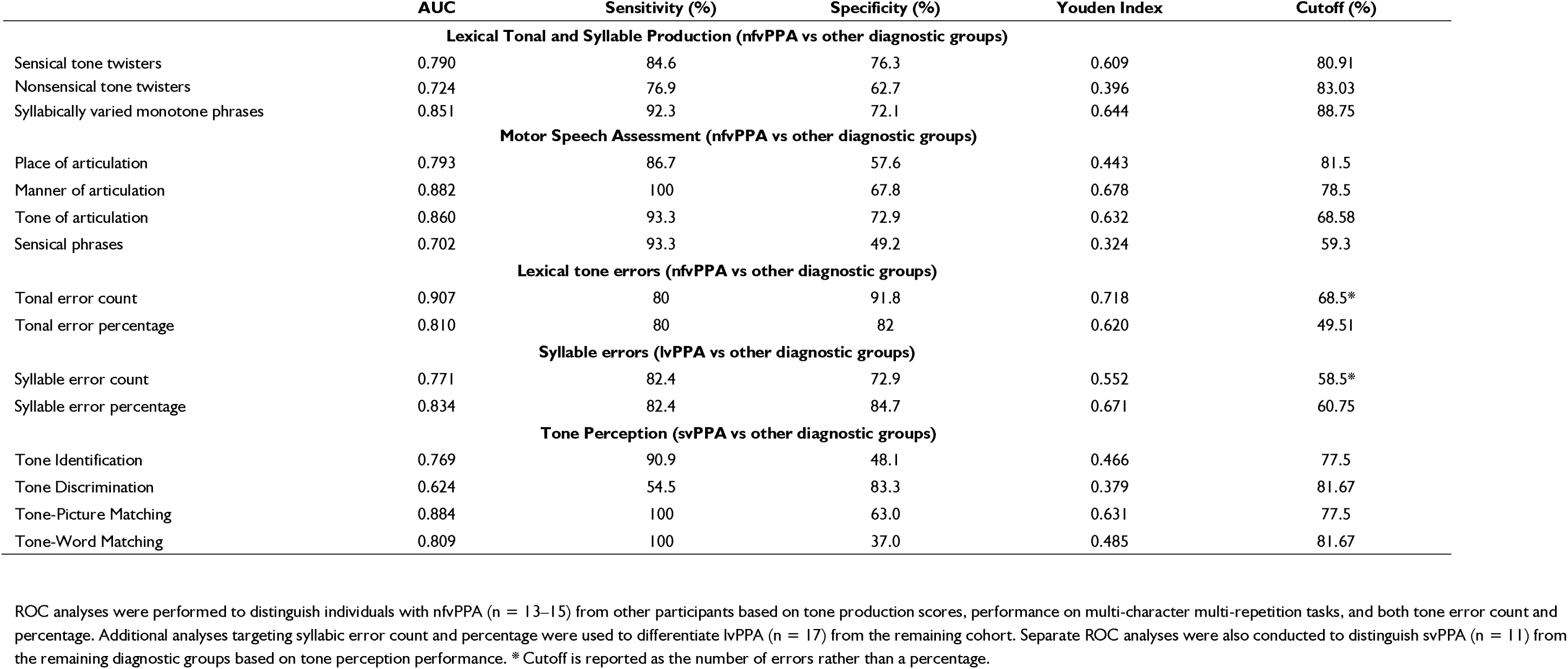
Receiver Operating Characteristic (ROC) Analysis of Lexical Tone Production and Perception Tasks.

In tone perception and comprehension tasks, the tone discrimination performance was associated with greater gray matter volume in the left superior, middle, and inferior temporal gyri, specifically in the middle and posterior temporal regions. Performance of tone identification test was positively associated with gray matter volume in the left orbitofrontal, anterior superior, middle, and inferior temporal gyri, insula and temporal pole. For tone-picture matching, the neural correlation analyses were observed in the left temporal pole, anterior temporal and fusiform gyri. Similarly, the score of tone-word matching test was positively associated the left temporal pole, anterior temporal, anterior cingulate, and medial superior frontal cortices.

## Discussion

This study illustrated one of the first in-depth investigation of lexical tone production and perception presentations in speakers of tonal languages with PPA, offering novel insight into the clinical phenotypes of PPA syndromes in this linguistically understudied population. In summary, Chinese-speaking nfvPPA participants exhibited significant impairments in lexical tone production, with a disproportionately high rate of tonal errors relative to syllabic errors. These findings suggest that impairments in lexical tone production may offer a clinically informative indicator into motor speech impairment in PPA. In contrast, Chinese-speaking svPPA patients showed pronounced deficits across several lexical tone perception tasks, underscoring the essential role of semantic representations in decoding pitch-based contrasts at the lexical level. These findings illuminate the importance of incorporating lexical tone assessments in the evaluation of tonal language speakers with PPA and provide a counterpoint to the universality of current PPA diagnostic frameworks, which has been largely informed by Indo-European linguistic paradigms.

### Lexical Tone Production in PPA and its neural signatures

Consistent with our hypothesis, Chinese-speaking individuals with nfvPPA demonstrated significant impairments in lexical tone production tasks, even after adjusting for age, education, and testing language. Notably, this deficit was apparent with the production of only two tone-twister phrases, underscoring the scalability and time efficiency of such assessment for clinical settings. Prior studies involving individuals with unilateral vocal fold paralysis, as well as brain tumor patients undergoing awake neurosurgery with electrocortical recordings, have highlighted that the production of lexical tone depends on precise and temporally synchronized control of the laryngeal and vocal fold musculature.^16,32^ We therefore posit that the tone production impairments observed in Chinese-speaking nfvPPA patients reflect disrupted coordination between the laryngeal articulatory motor systems. This interpretation is supported by converging neural correlations across both tone twister tasks in the left anterior insula and pars opercularis, regions critically involved in speech planning, and frequently implicated in AOS.^27,33–35^ The absence of primary motor cortex involvement further suggests such deficits are more primarily attributable to disruptions in motor programming than to impairments in muscular activation.

While both sensical and nonsensical tone twister tasks assess lexical tone production ability, our findings indicated that the nonsensical tone twisters demonstrated stronger associations with left ventral temporal regions. Given that stimuli were presented in both auditory and visual modalities, participants could engage either repetition or reading pathways, while converging on a shared cognitive mechanism for verbal production. We deduced sensical phrases places greater demands on the auditory stimuli and rely on the processes involving auditory-motor mapping and verbal working memory, both previously linked to lexical tone production.^36–38^ Conversely, in the absence of phrasal semantic coherence, participants appear to rely more heavily on orthographic decoding during the production of nonsensical tone twisters^39,40^, potentially accounting for the stronger associations observed with ventral temporal regions. These distinct processing demands account for the partially divergent neural correlates.^41^ Naturally, individuals with svPPA performed worse on nonsensical tone twister tasks, consistent with their ventral temporal degeneration affecting orthographic long-term memory.

Individuals with nfvPPA also demonstrated difficulties when producing monotone phrases composed of varied syllables, despite the consistency of lexical tone. We speculate this reflects motoric challenges in syllable articulation. Interestingly, performance on the monotone syllabic phrases strongly correlated with that on the tone twister tasks, suggesting that difficulties in both tone and syllable articulation may arise from shared underlying neural mechanisms.

### Motor Speech impairments and Apraxia of Speech in Chinese

AOS, a characteristics motor speech impairment in English-speaking nfvPPA patients, has been described differently in children across languages, including Arabic, Cantonese, and French.^42–45^ Specifically, Cantonese-speaking children with AOS were reported to lack features typically reported in English, such as voicing errors, lexical stress errors, and intrusive schwa insertions. Instead, they display difficulties with tone production and a tendency toward deaspiration.^46^ These cross-linguistic differences are further compounded by the inapplicability of standard English-language elicitation methods—relying on multisyllabic words and complex consonant clusters—to Chinese, which is predominantly monosyllabic and lacks consonant clusters.^27,47^ Consequently, linguistically tailored elicitation strategies are required when assessing Chinese-speaking individuals.

One of the earliest reports of lexical tone production deficits in a Cantonese-speaking adult with nfvPPA and AOS was by Tee et al.,^48^ with subsequent studies by Wong and colleagues demonstrating tone production impairments in Cantonese-speaking adults with post-stroke aphasia and AOS. ^49,50^ Collectively, these findings motivated the present study to examine the diagnostic utility of lexical tone as a strategy for eliciting motor speech impairment in Chinese-speaking individuals with PPA. Our results indicated that individuals with nfvPPA exhibited significantly reduced accuracy when repeating phrases that varied not only in place and manner of articulation, but also in lexical tone. These findings underscore the potential of lexical tone as a linguistically tailored strategy for eliciting AOS in Chinese speakers. Notably, nfvPPA patients produced both the greatest number and highest proportion of tonal errors relative to syllabic errors, a pattern not observed in other PPA variants, further reinforces the clinical relevance of lexical tone production in assessing motor speech function and highlights its potential as a diagnostic marker in tonal language populations.

While motor speech dysfunction is not a defining feature of lvPPA, lvPPA patients also demonstrated reduced accuracy on tone twister tasks relative to controls, though their performance surpassed that of the nfvPPA group. Nevertheless, error analyses revealed a predominance of syllabic rather than tonal errors in lvPPA, in contrast to the tonal error profile observed in nfvPPA. We speculate that while individuals with lvPPA may retain relatively intact motor speech planning abilities compared to nfvPPA, their increased syllabic errors likely reflect additional deficits in phonological processing.^51,52^

Lexical tones are generally classified into two main types: level tones, which maintain a consistent pitch, and contour tones, which exhibit a change in pitch over the course of the tone. Mandarin includes four tones: one level and three contour tones. In contrast, Cantonese distinguishes six tones: three level and three contour, with three additional checked level tones.^28^ (Supplement Figure 1) Intriguingly, only tone of articulation phrases showed a significant effect of testing language: Mandarin speakers outperformed Cantonese speakers. We attributed this to linguistic differences in both tonal system: Cantonese features a greater number of tones and more intricate tonal contours than Mandarin.

### Lexical Tone Perception in PPA and its neural basis

In the tone identification test, participants judged whether the presented tones existed in their testing language. Performance was associated with grey matter volume in a left-lateralized network encompassing the orbitofrontal cortex, temporal pole, anterior superior, middle, and inferior temporal gyri, and fusiform gyrus. We propose that these regions support tone identification through the integration of auditory, tonology, and decision-making processes. Specifically, the superior and middle temporal gyri likely subserve early auditory and tonal perception^17,53,54^, while the anterior temporal and fusiform regions may support the representation and access of tone-related semantic knowledge.^55–57^ Left orbitofrontal cortex may be engaged in the decision-making processes required to assess the linguistic relevance of tone stimuli.^58,59^ The impaired performance observed in svPPA and lvPPA aligns with their characteristic neurodegenerative patterns involving the left anterior and posterior temporal regions, respectively.^60^

In the tone discrimination task, previous studies have highlighted associations with the bilateral superior temporal gyri, a finding corroborated by our results.^17,18,54^ Although superior temporal gyrus is not among the earliest or most prominently affected regions in PPA and thus offers limited diagnostic utility, these findings nonetheless reinforce its fundamental role in the auditory processing of lexical tone.

Conversely, the tone-word and tone-picture matching tasks required participants to match auditory stimuli with homophones differentiated solely by tone, presented in word and picture formats, respectively. Although all three noncanonical PPA variants performed worse than controls, impairment was most severe in svPPA patients. Performance on these tasks depends on the integrity of lexicosemantic knowledge of tone and executive processes necessary for semantic judgment.^61–63^ These cognitive demands likely underlie the observed associations with left anterior temporal lobe atrophy for both tests,^61,62^ and the involvement of the anterior cingulate cortex in the tone-word matching task, consistent with previous findings linking this region to lexical tone reading.^63^ Additionally, the identification of tone perception impairments in Chinese PPA suggests that auditory comprehension difficulties in tonal language speakers may, in part, stem from deficits in lexical tone processing.

### Clinical significance of Lexical Tone in PPA

While these lexical tone and syllable production, and motor speech assessment task demonstrated strong AUC values for identifying nfvPPA, most exhibited high sensitivity with moderate specificity, with the exception of producing sensical tone twisters, tone of articulation and syllabically varied monotone phrases. Notably, the strong discriminant performance of the sensical tone twister and tone of articulation tests underscores the critical role of lexical tone in detecting motor speech impairment in Chinese-speaking nfvPPA patients. This is further substantiated by the high specificity of tone error frequency and count in distinguishing nfvPPA. Our findings highlight lexical tone production as a linguistically relevant and ecologically valid approach for assessing motor speech deficits in Chinese. Conversely, syllable error frequency and count were particularly elevated in lvPPA patients, demonstrating optimal sensitivity and specificity. Given that Chinese lacks nonsensical and sublexical phonological units for assessing phonological function, monitoring syllable errors in verbal production potentially serve as an important indicator of phonological deficits.

Lexical tone perception tasks, except for tone discrimination test, demonstrated high AUC values and sensitivity for identifying svPPA. These findings highlight the essential role of intact semantic knowledge in identifying pitch variation as lexical tone and mapping tones to their lexicosemantic representations. This pattern is conceptually analogous to reports of lexical stress assignment errors observed in Indo-European speakers with svPPA.^6–8^ In tonal languages, the distinction is more fine-grained and central to auditory comprehension. Nevertheless, the specificity of these tasks for detecting svPPA remains suboptimal, likely due to the involvement of additional cognitive mechanisms such as semantic judgment and auditory tone processing. Overall, our finding suggest that tone perception tasks may serve as a clinically useful screening tool for identifying svPPA in tonal language populations.

Clinical knowledge on PPA has largely been informed by studies involving speakers of Germanic and Romance languages.^64^ Given the existence of over 7,000 languages worldwide^65^, it is imperative to exercise caution when generalizing these findings, especially across languages that possess varying linguistic features compared to Indo-European languages. Our findings underline the heterogeneity of motor speech error profiles across PPA syndromes, challenge the adequacy of existing models based primarily on Indo-European languages and drawing attention to important diagnostic blind spots when non-English language speakers are assessed with verbatim translated tools. It is essential to incorporate additional linguistic components in a typologically informed manner, both to refine diagnostic precision and to guide the development of individualized speech therapy interventions that support the preservation of speech and language functions in PPA.

### Limitations and future directions

While this study offers important new insights, several limitations warrant consideration. First, our sample size, especially for imaging analyses, is comparable to previous PPA research, it nonetheless constrains the generalizability and statistical power of our findings. Second, given the varying lexical tone profiles of Mandarin and Cantonese, future studies should further stratify participants by Chinese language to elucidate how language-specific tonal systems influence neural engagement. Additionally, although tone and syllable error judgments were made by two formally trained native speakers, the manual nature of this process may limit its clinical applicability. Integrating machine learning approaches could help automate and scale this method for broader clinical use. Finally, longitudinal research is needed to assess the progression of tonal deficits over time and to determine their prognostic value in clinical staging.

## Conclusion

In summary, this study demonstrates that lexical tone processing is differentially impaired across PPA variants and that these impairments map onto distinct neural substrates. By integrating structural neuroimaging analysis, we offer new insights into the language–brain relationship in tonal speakers with neurodegenerative disease. These findings pave the way for more equitable and accurate diagnosis of PPA in linguistically diverse populations and underline the importance of including tone-based measures in future cross-linguistic PPA research.

## Acknowledgements

We would like to thank the participants and their families for their generous contributions to this work. We are also grateful to Stephanie Kwan, GuanXue Chen, Xin Wen, Anna Yao, and Kassey Chang at UCSF; Clara Cheng, Tammy Lau, and Kerwin Cheung in Hong Kong; as well as Yu-Ruei Lin, Yun-Ching Lo and Yu-Chung in Taiwan for their invaluable assistance with data collection and entry.

## Supplementary material

Supplementary material is available at *Brain* online.

