## Supplement Table 1 and Figure 1 and 2 for "Speaking in Tones: The role of lexical tones in Chinese-speaking Primary Progressive Aphasia"

**Supplement Table 1 The list of stimuli used on tone production tests.**

| Mandarin | Cantonese |
|---|---|
| **Sensical homophonous tonal phrases** | |
| 石 室 詩 士 施 氏，嗜 獅，誓 食 十 獅。 | 圓 圓 遠 遠 叫 圓 月。 |
| /shi2 shi4 shi1 shi4 shi1 shi4, shi4 shi1, shi4 shi2 shi2 shi1/ | /yun4 yun4 yun5 yun5 giu3 yun4 yut6/ |
| 媽 媽 騎 馬，馬 慢， 媽 媽 罵 馬。 | 施 氏 視 是 十 獅 逝 世。 |
| /ma1 ma1 qi2 ma3, ma3 man4, ma1 ma1 ma4 ma3/ | /si1 si6 si6 si6 sap6 si1 sai6 sai3/ |
| **Nonsensical homophonous tonal phrases** | |
| 媽 /ma1/ 馬/ma3/ 罵/ma4/ 麻/ma2/ | 因/jan1/ 忍/jan2/ 印/jan3/ 人/jan4/ 引/ jan5/ 刃 /jan6/ 壹/jat1/ 日/jat6/ |
| 達/da2/ 大/da4/ 打/da3/ 搭/da1/ | 分/fan1/ 粉/fan2/ 訓/fan3/ 焚/fan4/ 憤/fan5/ 份/fan6/ 忽/fat1/ 佛/fat6/ |
| 個/ge4/ 歌/ge1/ 隔/ge2/ 革/ge2/ | 空/hung1/ 紅/hung4/ 恐/hung2/ 控/hung3/ 虹/hung4/ |
| 韓/han2/ 汗/han4/ 喊/han3/ 鼾/han1/ | 施/si1/ 氏/si6/ 市/si5/ 時/si4/ 史/si2 事/si6/ |
| 看/kan4/ 刊/kan1/ 砍/kan3/ | 衣/yi1/ 椅/yi2/ 意/yi3/兒/yi4/ 耳/yi5/ 二/yi6/ |
| 人/ren2/ 忍/ren3/ 任/ren4/ | 淹/yim1/ 掩/yim2/ 厭/yim3/ 炎/yim4/ 染/yim5/ 驗/yim6/ |
| 扛/kang2/ 康/kang1/ 抗/kang4/ | |
| **Isotonous syllabically varied phrases** | |
| 風/feng1/ 淹/yan1/ 鄉村/xiang1 cun1/ | 風/fung1/ 淹/yim1/ 鄉村/heung1 chyun1/ |
| 人/ren2/ 遊/you2/ 河南/he2 nan2/ | 人/yan4/ 遊/yau4/ 河南/ho4 naam4/ |
| 粉/fen3/ 寫/xie/3 左手*/zuo3 shou3/ | 粉/fan2/ 寫/se2/ 左手/jo2 sau2/ |
| 字/zi4/ 驗/yan4/ 內陸/nei4 lu4/ | 字/ji6/ 驗/yim6/內陸/noi6 luk6/ |
| 尺/chi3/ 傻/sha3/ 剪紙*/jian3 zhi3/ | 尺/chek3/剎/chaat3/見證/gin3 jing3/ |
| 哨/shao4/ 是/shi4/ 見證/jian4 zheng4/ | 哨/saau3/ 試/si3/ 世界/sai3 gaai3/ |
| 母/mu3/ 買/mai3/ 美女*/mei3 nü3/ | 母/mou5/ 買/maai5/ 婦女/fu5 neui5/ |
| 熱/re4/ 夜/ye4/ 日月/ri4 yue4/ | 佛/fat6/ 服/fuk6/ 日月/yat6 yut6/ |
| 佛/fo2/ 福/fu2/ 從容/cong2 rong2/ | 屋/uk1/ 速/chuk1/ 即刻/jik1 hak1/ |
| 威/wei1/詩/shi1/ 揭發/ jie1 fa1/ | 威/wai1/ 詩/si1/ 聲音/sing1 yam1/ |
| **Multi-Character Multi-Repetition: Place of articulation** | |
| 偏偏天空 /pian1 pian1 tian1 kong1/ | 八撻嫁霸 /baat3 taat3 gaa3 baa3/ |
| 寶島港島/bao3 dao3 gang3 dao3/ | 答架八駕 /daap3 gaa3 baat3 gaa3/ |
| 販售現畫 /fan4 shou4 xian4 hua4/ | 趴卡潘他 /paa1 kaa1 pun1 taa1/ |

傳承成全 /chuan2 cheng2 cheng2 quan2/

贊助政見/zan4 zhu4 zheng4 jian4/

逼的即吉 /bik1 dik1 zik1 gat1/

司空西刊 /si1 hung1 sai1 hon1/

**Multi-Character Multi-Repetition: Manner of articulation**

密報破費 /mi4 bao4 po4 fei4/

內鬥測力 /nei4 dou4 ce4 li4/

主場首軟 /zhu3 chang3 shou3 ruan3/

新興金圈 /xin1 xing1 jin1 quan1/

觀光開花 /guan1 guang1 kai1 hua1/

哥苛刻娃 /go1 ho1 hak1 waa1/

瓜鴉哈跨 /gwaa1 aa1 haa1 kwaa1/

花爸媽趴 /faa1 baa1 maa1 paa1/

咱岔沙他 /zaa1 caa3 saa1 taa1/

啦哪沙渣 /laa1 naa1 saa1 zaa1/

**Multi-Character Multi-Repetition: Tone of articulation**

事實始失 /shi4 shi2 shi3 shi1/

麻媽罵馬 /ma2 ma1 ma4 ma3/

舅舅集九鳩 /jiu4 jiu ji2 jiu3 jiu1/

親情請求 /qin1 qing2 qing3 qiu2/

偉威圍魏 /wei3 wei1 wei2 wei4/

意兒宜衣 /ji3 ji4 ji4 ji1/

霞夏瑕哈 /haa4 haa6 haa4 haa1/

引孕忍印 /jan5 jan6 jan2 jan3/

意耳義爾 /ji3 ji5 ji6 ji5/

炎掩嚴染 /jim4 jim2 jim4 jim5/

**Multi-Character Multi-Repetition: Sensical phrases**

液晶電視螢幕 /ye4 jing1 dian4 shi4 ying2 mu4/

防彈背心 /fang2 dan4 bei4 xin1/

高速鐵軌 /gao1 su4 tie3 gui3/

通訊地址 /tong1 xun4 di4 zhi3/

大男人主義 /da4 nan2 ren2 zhu3 yi4/

鋼彈吊單槓 /gang1 dan4 diao4 dan1 gang4/

光芒萬丈 /guang1 mang2 wan4 zhang4/

初四吃素 /chu1 si4 shi1 su4/

黑肥發揮 /hei1 fei2 fa1 hui1/

龍惱農怒 /long2 nao3 nong2 nu4/

莫名奇妙 /mok6 ming4 kei4 miu6/

異曲同工 /ji6 kuk1 tung4 gung1/

膽識過人 /daam2 sik1 gwo3 jan4/

不相伯仲 /bat1 soeng1 baak3 zung6/

通訊地址 /tung1 seon3 dei6 zi2/

**Multi-Character Multi-Repetition: Nonsensical phrases**

牙威大紫 /nga4 wai1 daai6 ji2/

眼踏康島 /ngaan5 daap6 hong1 dou2/

泰銀及寒 /taai3 ngan2 kap6 hon4/

緣北紙次 /yun4 bak1 ji2 chi3/

句康牙本 /geui3 hong1 nga4 bun2/

Tables should be in an editable format, with cells clearly marked. Tab separated tables should not be used.
[a] Significant between Control and svPPA; [b] Significant between Control and nfvPPA; [c] Significant between Control and lvPPA; [d] Significant between svPPA and nfvPPA; e Significant between svPPA and lvPPA; [f] Significant between nfvPPA and lvPPA
[b]This is another footnote.

**Supplement Figure 1 The list of stimuli used on tone production tests.**

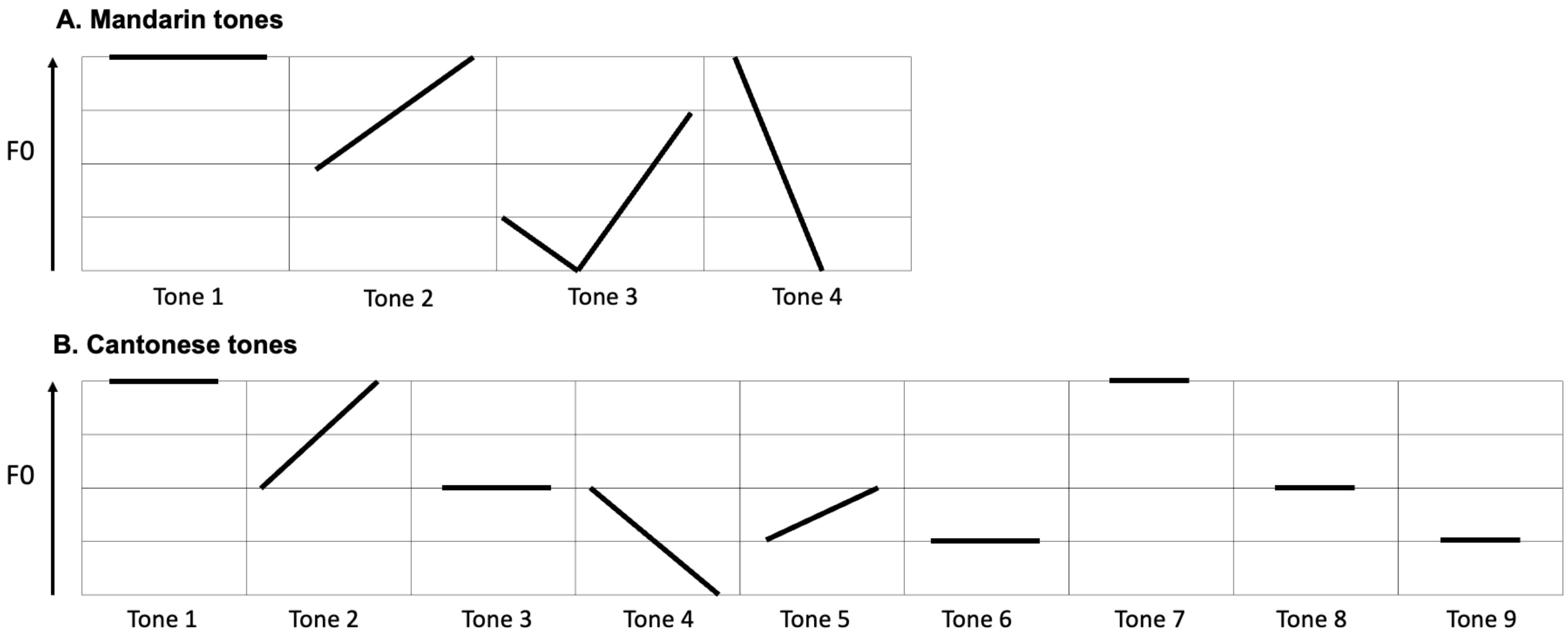

A. Mandarin tones
F0
Tone 1
Tone 2
Tone 3
Tone 4
B. Cantonese tones
F0
Tone 1
Tone 2
Tone 3
Tone 4
Tone 5
Tone 6
Tone 7
Tone 8
Tone 9

Supplement Figure 2. Cortical atrophy patterns across Primary Progressive Aphasia (PPA) subtypes

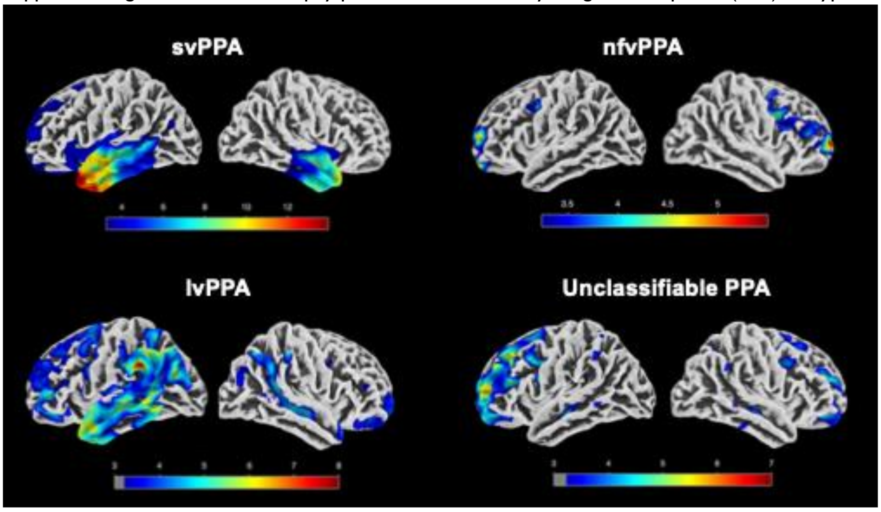


Surface-based statistical maps illustrate regions of significant cortical thinning in individuals from each Primary Progressive Aphasia (PPA) groups relative to cognitively normal controls enrolled in the CLAP study. Results are presented for four diagnostic groups: semantic variant PPA (svPPA, n = 10), nonfluent/agrammatic variant PPA (nfvPPA, n = 13), logopenic variant PPA (lvPPA, n = 15), and unclassifiable PPA (n = 10). Group comparisons were conducted using surface-based linear models implemented in the Computational Anatomy Toolbox (CAT12) within the Statistical Parametric Mapping (SPM) software, running under MATLAB R2022b. Analyses were adjusted for age, sex, and total intracranial volume. Color-coded statistical maps display *t*-values derived from vertex-wise group comparisons, with brighter intensities indicating greater cortical thinning in patients relative to cognitively normal controls. Results are thresholded at $p < 0.001$ (uncorrected) with a minimum cluster size of $k > 100$ vertices.
